# Relationships of residential distance to Greenhouse Floriculture and Organophosphate, Pyrethroid, and Neonicotinoid Urinary Metabolite Concentration in Ecuadorian Adolescents

**DOI:** 10.1101/2024.10.23.24315947

**Authors:** Briana N.C. Chronister, Georgia L. Kayser, Franklin de la Cruz, Jose Suarez-Torres, Dolores Lopez-Paredes, Sheila Gahagan, Harvey Checkoway, Marta M. Jankowska, Jose Ricardo Suarez-Lopez

## Abstract

**Background:** Adolescents living in agricultural areas are at higher risk of secondary pesticide exposure; however, there is limited evidence to confirm exposure by pesticide drift for greenhouse floriculture, like rose production.

**Methods:** 525 adolescents (12-17, 49% male) living in Pedro Moncayo, Ecuador were assessed in 2016. Urinary concentrations of creatinine and pesticide biomarkers (organophosphates, neonicotinoids, and pyrethroids) were measured using mass-spectrometry. Home distance to the nearest greenhouse and surface area of greenhouses within various buffer sizes around the home were calculated. Linear regression assessed whether home distance and surface area of greenhouses was associated with creatinine-adjusted metabolite concentration, adjusting for demographic, socioeconomic, and anthropometric variables. Geospatially weighted regression (GWR) was conducted, adjusting for similar covariates. Getis-ord Gi* identified hot and cold spots using a 1994m distance band.

**Results:** The associations between residential distance to greenhouses and urinary pesticide metabolites differed by metabolite type. The adjusted mean concentrations of OHIM (neonicotinoid) were greater (p-difference=0.02) among participants living within 200m (1.08 ug/g of creatinine) vs >200m (0.64 ug/g); however, the opposite was observed for 3,5,6-Trichloro-2-pyridinol (TCPy, organophosphate; 0-200m: 3.63 ug/g vs >200m: 4.30 ug/g, p-diff= 0.05). In linear models, greater distances were negatively associated with para-nitrophenol (PNP, organophosphate; percent difference per 50% greater distance [95% CI]: -2.5% [-4.9%, -0.1%]) and somewhat with 2-isopropyl-4-methyl-6-hydroxypyrimidine (IMPy, organophosphate; -4.0% [-8.3%, 0.4%]), among participants living within 200m of greenhouses. Concurring with the adjusted means analyses, opposite (positive) associations were observed for TCPy (2.1% [95%CI: 0.3%, 3.9%]). Organophosphate and pyrethroid hotspots were found in parishes with greater greenhouse density, whereas neonicotinoid hot spots were in parishes with the lowest greenhouse density.

**Conclusion:** We observed negative associations between residential distance to greenhouses with OHIM, PNP and to some extent IMPy, suggesting that imidacloprid, parathion and diazinon is drifting from floricultural greenhouses and reaching children living within 200m. Positive TCPy associations suggest greenhouses weren’t the chlorpyrifos source during this study period, which implies that non-floricultural open-air agriculture (e.g. corn, potatoes, strawberries, grains) may be a source. Further research incorporating diverse geospatial constructs of pesticide sources, pesticide use reports (if available), participant location tracking, and repeated metabolite measurements is recommended.

## 1. Introduction

Children living in agricultural communities are at higher risk of para-occupational pesticide exposure due to pesticide drift from spray sites to non-targeted regions and nearby homes, primarily in the form of droplets or evaporation.^1–5^ Pesticide exposure through pesticide drift has been found to decay with distance; hence, individuals living closer to agricultural fields are potentially at higher risk of exposure. In Eastern Washington, United States, when comparing household dust and soil from homes within 61 meters (200 ft) versus a 402 m (1320 ft) away from an agricultural field,^6^ homes closer to agricultural fields had significantly higher concentrations of organophosphates detected in dust and soil samples.^6^ Another study in Eastern Washington found that a 1609 meter (one mile) increase in home distance from an agricultural field resulted in a 20% decrease in dimethylphosphate urinary metabolite concentrations in a sample of 100 farm workers and 100 non-farm working adults and children.^7^ A study of households located in an agricultural community in North Carolina and Virginia identified that homes located next to an agricultural field had 18 times higher odds of detectable levels of pesticides in household dust, compared to household that were not next to a field.^8^ Some pesticides that were measured include carbamates (e.g., carbaryl), organochlorines (e.g., heptachlor), organophosphates (e.g., diazinon, disulfoton), herbicides (e.g., oxyfluorfen, pendimethalin), and pyrethroids (e.g., *cis-*permethrin, *trans*-permethrin).^9^ While a study conducted in rural Taiwan found that carbofuran and tetramethrin in house dust were significantly correlated with rice cultivation area at 50m, 150m, and 200m buffer distances.^10^

Pesticide drift from open-air agriculture (e.g., corn production) has been characterized in the literature, but few studies have described the relationship between proximity to greenhouse agriculture (e.g., rose production) and individual pesticide exposure patterns. In one study, environmental pesticide distribution was examined for small-scale floricultural greenhouses where backpack sprayers were used to apply chlorothalonil to seedlings of geranium, fuchsia and balsam.^11^ Of the total volume of pesticides applied, 0.70% - 19.5% was detected on the greenhouse walls, 2.3 - 51.9% was found in the soil, and 84.1% - 96.2% was found on the crops.^11^ Personal air sampling of greenhouse workers found respiratory exposure from 4.5% to 49.5%, dependent on organophosphate assessed, demonstrating its potential to be volatilized.^12^

Although pesticides are developed through strict regulations to maximize effectiveness while minimizing impacts on human health,^13^ researchers have found negative health outcomes that stem from pesticide exposure, not specific to certain classifications.^14–18^ In adults these include acute health effects, such as irritation, weakness or fatigue, and changes in mood, to chronic health effects, such as neurobehavioral changes, hypertension, diabetes, endocrine disruption, and leukemia.^19–23^ Closer proximity to agricultural fields not only increases the pesticide exposure potential ^24^, but may increase related health risks such as developmental delays,^25^ lower neurobehavioral performance,^25,26^ and autism^25^ in children. Adverse health consequences in children have been consistently observed with exposures to insecticides including organophosphates and pyrethroids, and to a lesser extent with neonicotinoids. Organophosphate and pyrethroid exposure in infancy or childhood has been linked to poorer neurobehavioral and cognitive performance.^27–33^ Studies of neonicotinoid exposure in mammals and humans suggest that they may lead to negative reproductive effects, neurotoxicity, and hepatotoxicity. ^34–37^

Due to its favorable climate, lower labor costs, and demand for cut flowers, the Ecuadorian Andes region has had an exponential increase in its floriculture industry since its inception in 1983, leading to an annual value of cut flower exports of 1.2 billion dollars.^38^ The main importers of flowers from Ecuador are the United States, Russia, and the Netherlands.^39,40^ Pedro Moncayo County in the province of Pichincha, Ecuador is one of the country’s epicenters of rose production.^41,42^ Roses in this region are grown in greenhouses, and studies of pesticide drift patterns among this type of agriculture method is limited compared to open field agriculture.^14^ Within the study of Secondary Exposures to Pesticides among Children and Adolescents (ESPINA: Estudio de la Exposicion Secundaria a Plaguicidas en Ninos y Adolescentes [Spanish]), we previously found that home proximity to floricultural greenhouses, especially within 275 m, was associated with lower acetylcholinesterase (AChE) activity among Ecuadorian children, which indicated greater exposure to cholinesterase inhibitor insecticides from off-target pesticide drift.^43^ Further work to characterize which specific pesticides may be reaching populations near floricultural greenhouses is needed. We also found that adolescents living within 100m of a floricultural greenhouse had increased odds of lower neurobehavioral performance.^26^ The aim of this study was to evaluate the association between residential proximity to greenhouse floriculture and urinary pesticide metabolites in adolescents living in agricultural settings. We hypothesize that greater residential distance to floriculture was negatively associated with urinary pesticide metabolite concentrations, while increasing floriculture surface area near participants’ homes will be positively associated with pesticide metabolite concentrations.

## 2. Methods

### Study site

Pedro Moncayo County, located in the Ecuadorian Andes, is composed of five parishes (Malchingui, Tocachi, La Esperanza, Tabacundo, and Tupigachi) and houses a large floriculture industry that is vital to the County’s economy (Fig. 1). In 2004 it was estimated that Floriculture employs 21% of all adults.^44^ While in 2016 greenhouse floricultural crop land comprised 4.47% (1495 ha) of the geographic area of the county^43,45^ , which doubled from 2008 where 2% of the county was comprised of floricultural greenhouses (656 ha).^43^ From East to West, the total surface area composed of floricultural greenhouses for each parish was 0.05% for Malchingui, 0.12% for Tocachi, 1.12% for La Esperanza, 13.1% for Tabacundo, and 12.4% for Tupigachi.

**Figure 1.**
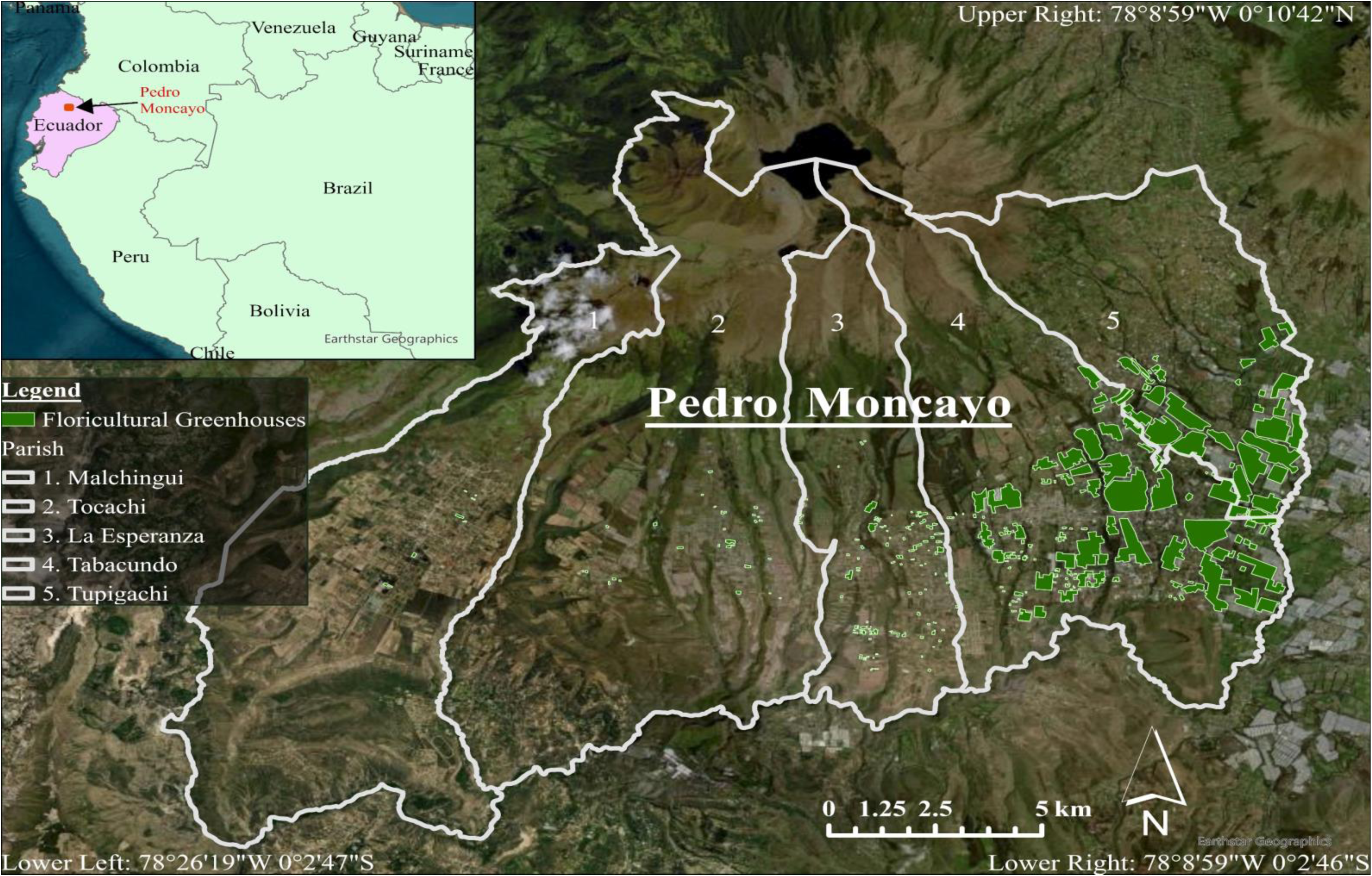
Study Region of Pedro Moncayo. The floricultural fields are those present in 2016. The image source is the ArcGIS base map with a contemporary image of the region.

Flower crops in Pedro Moncayo are sprayed with over 50 different fungicides and over 20 different insecticides by workers using hand sprayers. Greenhouse flower crops in Pedro Moncayo are sprayed with over 50 different fungicides and over 20 different insecticides by workers using hand sprayers.^46,47^

### Participants

A total of 313 boys and girls (4-9 years) were examined in 2008 as part of the ESPINA study; Study inclusion criteria for the 2008 cohort comprised the following: (1) children living with flower workers: cohabitation with a floricultural greenhouse worker for at least one year; (2) children living without agricultural workers: never cohabited with an agricultural worker, never inhabited a house where agricultural pesticides were stored and having no previous direct contact with pesticides. ^40,44^ Participants were identified and recruited using the Survey of Access and Demand of Health Services in Pedro Moncayo County. This study focuses on the follow-up examination of participants from July through October in 2016, which included 238 participants examined in 2008 and 297 new adolescent volunteers (n=535) of ages 11-17 years. Recruitment was conducted using the System of Local and Community Information (SILC). The SILC contains information of the 2016 Pedro Moncayo County Community Survey (formerly the Survey of Access and Demand of Health Services in Pedro Moncayo County) and was used to identify eligible participants for the study. Further details about participant recruitment have been published previously.^48,49^

*Ethical approval:* This study was approved by the institutional review boards at the University of California San Diego, Universidad San Francisco de Quito and the Ministry of Public Health of Ecuador. Informed consent from parents was obtained for participation in this study and parental permission of participation for each of their selected children was obtained. Child assent was obtained for all children.

### Data collection

Trained examiners were blinded to all participants’ pesticide exposure status and were unaware as to whether they did or did not live with a floricultural worker. Children’s height was measured following recommended procedures to the nearest 1 mm using a height board, and weight was measured using a digital scale (Tanita model 0108MC; Corporation of America, Arlington Heights, IL, USA). ^50^ Information on income, education level, and other demographic information was collected through home interviews with the participants’ parents.

In 2016, participants were asked to collect urine samples upon awakening and bring them to the examination site where the urine was aliquoted and frozen at -20° C. At the end of each day, samples were transported to Quito for storage at -80° C. Samples were then transported overnight to UCSD at -20° C using a courier and stored at -80° C at UCSD. Samples were then shipped overnight at -20° C from UCSD to the National Center for Environmental Health, Division of Laboratory Sciences of the CDC (Atlanta, GA) for quantification of all organophosphates, neonicotinoids, and pyrethroids, and to the Laboratory for Exposure Assessment and Development in Environmental Research at Emory University (Atlanta, GA) for quantification of creatinine.

### Pesticide Metabolite Quantification

A total of thirteen insecticide metabolites of organophosphates, pyrethroids, and neonicotinoids were measured once for each participant. This included six neonicotinoid metabolites (acetamiprid [ACET], acetamiprid-N-desmethyl [AND], clothianidin [CLOT], imidacloprid [IMID], 5-hydroxy imidacloprid [OHIM] and thiacloprid [THIA]), four organophosphates metabolites (2-isopropyl-4-methyl-6-hydroxypyrimidine [IMPy], 3,5,6-Trichloro-2-pyridinol [TCPy], para-Nitrophenol [PNP], and malathion dicarboxylic acid [MDA]), and three pyrethroid metabolites (3-phenoxybenzoic acid [3-PBA], 4-fluoro-3-phenoxybenzoic acid [4F-3PBA] and trans-3-(2,2-Dichlorovinyl)-2,2-dimethylcyclopropane carboxylic acid [*trans-DCCA]*). Metabolites were excluded from analysis if they had a limit of detection (LOD) below 15%, which included four neonicotinoid metabolites (ACET, CLOT, IMID, and THIA) and one pyrethroid metabolite (4F-3PBA).^46,47^

Targeted organophosphate and pyrethroid metabolites were quantified using liquid chromatography coupled with tandem mass spectrometry and isotope dilatation.^51^ The limit of detection (LOD) was 0.1 μg/L for TCPy, PNP, IMPy, and 3-PBA, 0.5 μg/L for MDA, and 0.6 μg/L for *trans*-DCCA. To measure neonicotinoid metabolite concentrations, targeted metabolites were quantified from enzymatic hydrolysis of 0.5 mL of urine and online solid phase extraction to release, extract and concentrate the target biomarkers, followed reversed-phase high-performance liquid chromatography-tandem mass spectrometry (HPLC-MS/MS) using electrospray ionization (ESI).^52^ The LOD was 0.2 μg/L for AND, and0.4 μg/L for OHIM. Pesticide metabolite concentrations below the LOD were imputed by diving the LOD by the square root of two. This was done for 62.3% of MDA, 75.8% of IMPy, 71.2% of OHIM, 62.2% of AND, 11.1% for 3-PBA, and 80.8% of *trans*-DCCA observations. Higher metabolite concentrations are indicative of having higher pesticide exposure.

To assess the dilution of urine, creatinine measurement was conducted and controlled for in the analysis. Urinary creatinine was quantified using HPLC-MS/MS with ESI.^53^ The LOD was 5 mg/dL with an RSD of 7%. All metabolite concentrations were divided by the creatinine concentration (g/dL) to control for urine dilution.^54,55^ Metabolites with values below the LOD were imputed as LOD/√2 for all analyses. Summary scores for each pesticide classification was created by natural log (ln) transforming the imputed creatinine adjusted metabolite concentrations plus 1, dividing it by the standard deviation, and then averaging all the metabolites of the same classification. One was added to the metabolites to ensure the summary score was above zero, which allowed us to ln-transform the variable. When individual metabolites were used in our models, the concentrations were ln-transformed without adding 1.

Quality control/quality assurance protocols were followed to ensure data accuracy and reliability of the analytical measurements. All the study samples were re-extracted if quality control failed the statistical evaluation. ^52,56^

*Geospatial Variables*. Geographical coordinates of participants’ homes were obtained through portable global positioning system receivers in 2016 using SILC data, led by the Fundación Cimas del Ecuador. Floricultural greenhouses(polygons) were created using satellite imagery from 2016. Country-level, state-level, and county-level borders were provided by the World Bank.^57^ Distance to the nearest floricultural greenhouses was calculated by assessing the distance in meters from homes to the nearest contour of greenhouses. Buffers around participants’ homes were created for 100m, 150m, 200m, 300m, 500m and 750m, and the surface area of floricultural greenhouses within each buffer was computed.^41^

*Statistical Analysis*. We calculated the means and standard deviation (SD) for age, BMI-for-age z-score, height-for-age z-score, parental education, geometric means and adjusted least square means (ls-means) for pesticide metabolites, geometric means for average monthly salary and pesticide metabolite concentrations, and proportions for each category of gender, race, and cohabitation with a flower or agricultural worker. We calculated the p-value for trend (p-trend) for participant characteristics across strata of those who did and did not live within 200m of the nearest greenhouse (200m strata); rationale for using this strata can be found below.

Generalized linear models (GLM) estimated the associations of distance from the home to the closest greenhouse (home distance) and greenhouse surface area within aforementioned buffers (100m-750m) with ln-transformed pesticide metabolite concentrations. GLM models of home distance and metabolite concentrations were stratified using a 200m cut point to assess threshold effects. The strata was determined after conducting sensitivity analysis with 100m, 150m, 200m, 250m, 300m, 400m, and 750m cut points (Table S1).Using the minimum p-value approach^58^, the 200m strata was chosen. The threshold is supported by prior studies which found lower AChE activity among children living within 150m to 275m of a greenhouse,^43^ and steep degradations of air contamination by pesticide application after 250m.^59^ As the dependent and independent variables were natural log (ln)-transformed, the models estimated the percent increase in pesticide metabolite concentration for every 50% increase of greenhouse surface area (m^2^) or distance (m) to the nearest greenhouse. This was done by calculating 1.5 to the power of the coefficient, subtracting 1, and multiplying by 100. All GLM models controlled for sex (female or male [reference]), age (years), race (‘Indigenous’ or ‘White or Mestizo’ [reference]), height-for-age z-score, BMI-for-age z-score, average monthly income (United States Dollar), average parental education (years) and whether the participant lived with an agricultural or floricultural worker (Yes or No [reference]). that take-home exposure (para-occupational) is another important source of secondary pesticide exposure, we controlled for this in the model to solely account for exposure to pesticides due to proximity.^60^ These covariates were determined *a priori* based on similar analyses in the literature ^43^. GLM model residuals were output to test spatial autocorrelation (see below).

Curvilinearity was assessed by introducing a squared term of home distance into the model and was reported if it was statistically significant (p<0.05). Locally weighted polynomial regression (LOESS) curve graphs were created to visualize statistically significant linear and curvilinear associations. The LOESS used fully adjusted least squares means of the creatinine adjusted imputed metabolite for 350 ranks of home distance to the nearest floricultural greenhouse, and were plotted with a 0.8 smoothness level.

The proportion of metabolite concentrations below the LOD exceeded 5-10% for all metabolites except PNP and TCPy, thus biased results may bias variance estimates. ^61^ To address this, Tobit regression was conducted to assess the association between home distance to the nearest floricultural greenhouse and floricultural greenhouses surface area within a 150m buffer with MDA, IMPy, OHIM, AND, 3-PBA, and *trans*-DCCA. This was done to determine whether the generalized linear models were biasing our results. Similar to the linear regression models, the exposures were ln-transformed and the metabolites were imputed with a constant, adjusted for creatinine concentration, and were log transformed. The Tobit model was censored using a lower bound based on the metabolite LOD (e.g. LOD for MDA was 0.5 µg/L) and transformed to match the metabolites. Thus, the lower bound was determined by dividing the LOD by creatinine and then ln-transforming it.

To assess special dependency of the linear regression model, Moran’s index (I) tests of spatial autocorrelation were conducted on output model residuals of the linear associations between home distance and biomarker concentrations. A statistically significant Moran’s I indicate that there is spatial clustering of errors, a potential indicator for model misspecification.^62^ If clustering of residuals was identified, we calculated the Getis-Ord Gi* statistic to identify hot and cold spots within a 90% confidence interval; golden search identified an ideal Euclidean distance band of 1994 m.^63,64^ To de-identify individuals’ homes, points with a non-significant Gi* statistic were masked, and hot and cold spot diameters were increased to 850 meters.

To identify spatial variation between the association of home distance to greenhouses and urinary metabolite concentration, we conducted geographically weighted regression (GWR). The GWR model creates a separate regression equation for each home, allowing the association to vary across the county for each location of interest (*i)*. This is denoted using the following equation.

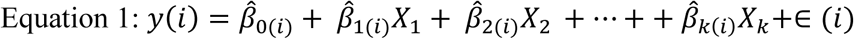

In this equation y(i) is the observation of the dependent variable at the ith location, *β̂*_0(*i*)_ is the estimated intercept at the *i*th location, Xk(i) is the observation of the kth explanatory variable at the *i*th location, *β̂_k_*_(*i*)_ is the kth parameter estimate at the *i*th location, and e(*i*) is the random error term for i ^65,66^. Parameter estimates are obtained at each location by calibrating a locally weighted regression using the following estimator:

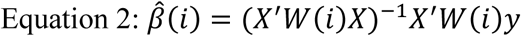

*β̂*(*i*) is an k ×1 vector of parameter estimates, X is an n × k matrix of explanatory variables, y is a k × 1 vector of observations for the dependent variable, and W (*i)* is a spatial weights matrix designed to allow data points closer to location *i* to have a stronger influence on the local regression ^65,66^.

GWR model used the golden search neighborhood selection method to identify the optimal distance band between 4 and 10 kilometers (km). The GWR assessed for the association between home distance to floricultural greenhouses with creatinine adjusted TCPy and PNP concentrations, adjusting for age, BMI for age z-score, average monthly salary, average parental education, and cohabitation with a floricultural or agricultural worker. To improve interpretation of the GWR estimates, home distance to the nearest agricultural field was divided by 500. Thus, the estimate represents the difference in metabolite concentration for a 500m change in home distance to the nearest agricultural field. The GWR 500m distance coefficient and standard errors were mapped, and each point was given a 1 km diameter to mask participants’ homes. All statistical analyses were conducted in SAS (Cary, NC) and geographic analyses were conducted in ArcGis Pro 2.8.0 (Esri Inc.).

## 3. Results

Of the 535 participants who were assessed in 2016, 525 participants were measured for organophosphate, neonicotinoid, and pyrethroid metabolites and were included in the analysis. The mean age of the participants was 14.43 years (SD=1.76) (Table 1). The average height-for-age z-score was -1.49 (SD=0.86), while the average BMI-for-age z-score was 0.39 (SD=0.86) indicating that participants were short in stature but with normal BMI (Table 1). Individuals who lived within 200m of the nearest greenhouse were more likely to be Mestizo or White and have higher creatinine concentration (Table 1). The geometric means of all pesticide urinary metabolites observed in the cohort are listed in Table 1.

**Table 1.**
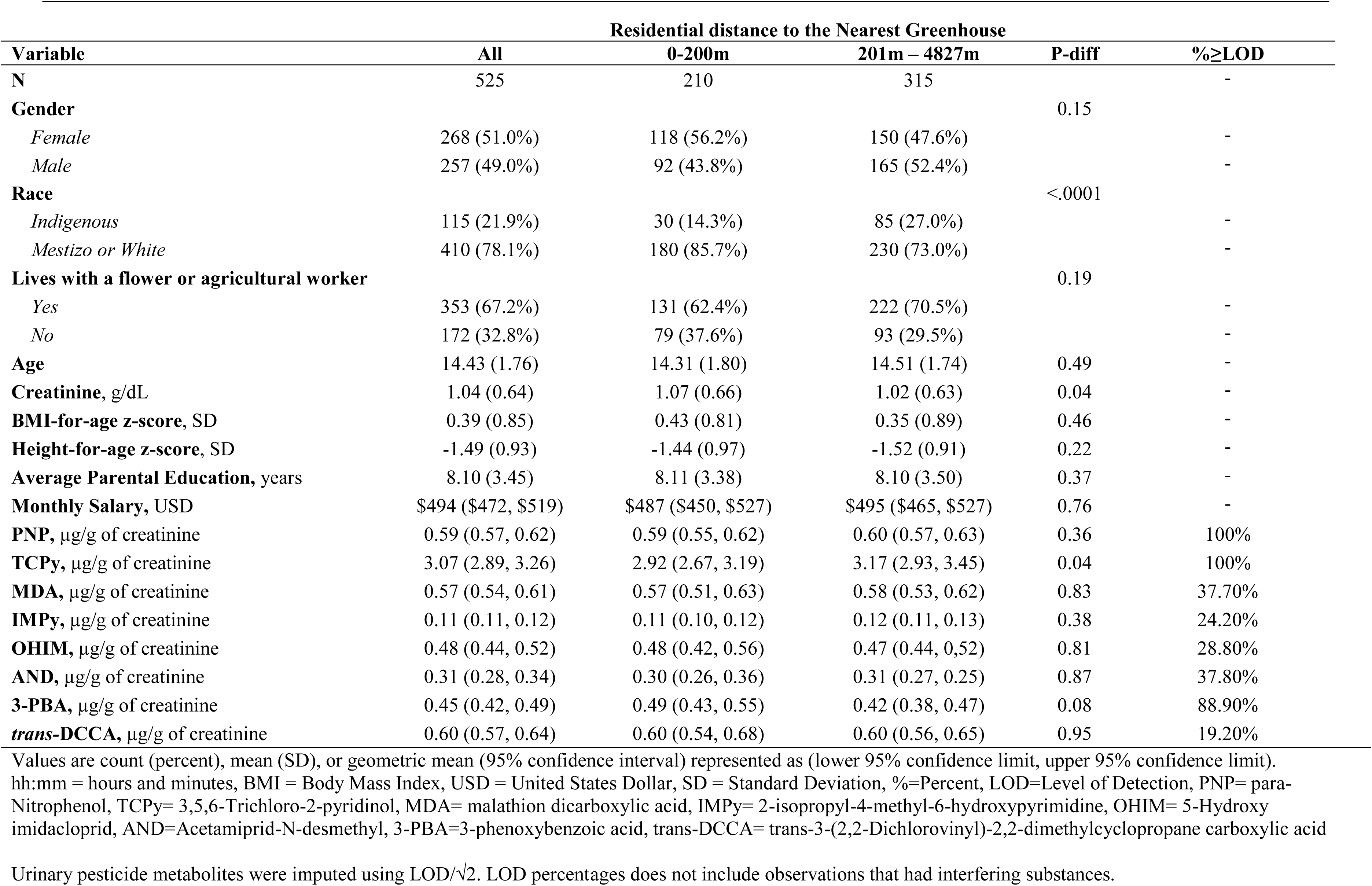
Participant characteristics stratified by residential distance to the nearest greenhouse.

| Variable | Residential distance to the Nearest Greenhouse |  |  | P-diff | %≥LOD |
| --- | --- | --- | --- | --- | --- |
|  | All | 0-200m | 201m – 4827m |  |  |
| N | 525 | 210 | 315 |  | - |
| **Gender** |  |  |  | 0.15 |  |
| *Female* | 268 (51.0%) | 118 (56.2%) | 150 (47.6%) |  | - |
| *Male* | 257 (49.0%) | 92 (43.8%) | 165 (52.4%) |  | - |
| **Race** |  |  |  | <.0001 |  |
| *Indigenous* | 115 (21.9%) | 30 (14.3%) | 85 (27.0%) |  | - |
| *Mestizo or White* | 410 (78.1%) | 180 (85.7%) | 230 (73.0%) |  | - |
| **Lives with a flower or agricultural worker** |  |  |  | 0.19 |  |
| *Yes* | 353 (67.2%) | 131 (62.4%) | 222 (70.5%) |  | - |
| *No* | 172 (32.8%) | 79 (37.6%) | 93 (29.5%) |  | - |
| **Age** | 14.43 (1.76) | 14.31 (1.80) | 14.51 (1.74) | 0.49 | - |
| **Creatinine, g/dL** | 1.04 (0.64) | 1.07 (0.66) | 1.02 (0.63) | 0.04 | - |
| **BMI-for-age z-score, SD** | 0.39 (0.85) | 0.43 (0.81) | 0.35 (0.89) | 0.46 | - |
| **Height-for-age z-score, SD** | -1.49 (0.93) | -1.44 (0.97) | -1.52 (0.91) | 0.22 | - |
| **Average Parental Education, years** | 8.10 (3.45) | 8.11 (3.38) | 8.10 (3.50) | 0.37 | - |
| **Monthly Salary, USD** | \$494 (\$472, \$519) | \$487 (\$450, \$527) | \$495 (\$465, \$527) | 0.76 | - |
| **PNP, µg/g of creatinine** | 0.59 (0.57, 0.62) | 0.59 (0.55, 0.62) | 0.60 (0.57, 0.63) | 0.36 | 100% |
| **TCPy, µg/g of creatinine** | 3.07 (2.89, 3.26) | 2.92 (2.67, 3.19) | 3.17 (2.93, 3.45) | 0.04 | 100% |
| **MDA, µg/g of creatinine** | 0.57 (0.54, 0.61) | 0.57 (0.51, 0.63) | 0.58 (0.53, 0.62) | 0.83 | 37.70% |
| **IMPy, µg/g of creatinine** | 0.11 (0.11, 0.12) | 0.11 (0.10, 0.12) | 0.12 (0.11, 0.13) | 0.38 | 24.20% |
| **OHIM, µg/g of creatinine** | 0.48 (0.44, 0.52) | 0.48 (0.42, 0.56) | 0.47 (0.44, 0.52) | 0.81 | 28.80% |
| **AND, µg/g of creatinine** | 0.31 (0.28, 0.34) | 0.30 (0.26, 0.36) | 0.31 (0.27, 0.25) | 0.87 | 37.80% |
| **3-PBA, µg/g of creatinine** | 0.45 (0.42, 0.49) | 0.49 (0.43, 0.55) | 0.42 (0.38, 0.47) | 0.08 | 88.90% |
| **trans-DCCA, µg/g of creatinine** | 0.60 (0.57, 0.64) | 0.60 (0.54, 0.68) | 0.60 (0.56, 0.65) | 0.95 | 19.20% |

Values are count (percent), mean (SD), or geometric mean (95% confidence interval) represented as (lower 95% confidence limit, upper 95% confidence limit). hh:mm = hours and minutes, BMI = Body Mass Index, USD = United States Dollar, SD = Standard Deviation, %=Percent, LOD=Level of Detection, PNP= para-Nitrophenol, TCPy= 3,5,6-Trichloro-2-pyridinol, MDA= malathion dicarboxylic acid, IMPy= 2-isopropyl-4-methyl-6-hydroxypyrimidine, OHIM= 5-Hydroxy imidacloprid, AND=Acetamiprid-N-desmethyl, 3-PBA=3-phenoxybenzoic acid, trans-DCCA= trans-3-(2,2-Dichlorovinyl)-2,2-dimethylcyclopropane carboxylic acid
Urinary pesticide metabolites were imputed using LOD/ $\sqrt{2}$ . LOD percentages does not include observations that had interfering substances.

### Home distance to floriculture greenhouses and urinary metabolite concentration

The adjusted ls-means for OHIM were higher (p=0.02) among participants who lived within 200m of a greenhouse (1.08 µg/g [95%CI: 0.80, 1.36]), compared to those who lived farther than 200m (0.64 µg/g [0.42, 0.87]) (Table 2). However, TCPy ls-mean concentrations were lower (p=0.05) among those who lived within 200m from the nearest greenhouse (3.63 µg/g [3.12, 4.14]) compared to those living beyond this distance (4.30 µg/g [0.88, 4.71]). Ls-mean differences for other metabolites were not statistically different between the groups.

**Table 2.** Least squares mean of organophosphate, neonicotinoid, and pyrethroid metabolites (µg/g of creatinine) stratified by home distance to the nearest greenhouse (0-200m to >200m).

| Adjusted Least Squares Means (95% CI) |  |  |  |  |  |
| --- | --- | --- | --- | --- | --- |
|  | All | 0-200m | 201m - 4827m | P-difference | %≥LOD |
| n | 525 | 210 | 315 |  |  |
| **PNP** | 0.67 (0.64, 0.70) | 0.67 (0.61, 0.70) | 0.67 (0.63, 0.71) | 0.63 | 100% |
| **TCPy** | 4.03 (3.71, 4.35) | 3.63 (3.12, 4.14) | 4.30 (0.88, 4.71) | 0.05 | 100% |
| **MDA** | 0.76 (0.69, 0.84) | 0.77 (0.65, 0.88) | 0.77 (0.67, 0.86) | 0.96 | 37.7% |
| **IMPy** | 0.19 (0.16, 0.22) | 0.17 (0.12, 0.22) | 0.20 (0.16, 0.25) | 0.36 | 24.2% |
| **OHIM** | 0.81 (0.64, 0.99) | 1.08 (0.80, 1.36) | 0.64 (0.42, 0.87) | 0.02 | 28.8% |
| **AND** | 0.80 (0.57, 1.03) | 0.83 (0.45, 1.21) | 0.78 (0.48, 1.08) | 0.86 | 37.8% |
| **3-PBA** | 0.71 (0.62, 0.80) | 0.76 (0.62, 0.91) | 0.67 (0.56, 0.79) | 0.33 | 88.9% |
| **trans-DCCA** | 0.83 (0.75, 0.92) | 0.89 (0.75, 1.03) | 0.79 (0.68, 0.90) | 0.28 | 19.2% |

Least squares mean were obtained by running a generalized linear regression model with the creatinine adjusted metabolite concentrations that were imputed with a constant as the dependent variable, and a categorical variable using a 200m split of the home distance to the nearest floricultural greenhouse as the independent variable. The model was adjusted for age, height-for-age z-score, BMI-for-age z-score, race, gender, monthly income, parental education, living with an agricultural or flower worker. P-value represent if the least squares means of the urinary metabolite concentrations were statistically significantly different across the two strata.
CI= Confidence Interval, %=Percent, LOD=Level of Detection, PNP= para-Nitrophenol, TCPy= 3,5,6-Trichloro-2-pyridinol, MDA= malathion dicarboxylic acid, IMPy= 2-isopropyl-4-methyl-6-hydroxypyrimidine, OHIM= 5-Hydroxy imidacloprid, AND=Acetamiprid-N-desmethyl, 3-PBA=3-phenoxybenzoic acid, trans-DCCA= trans-3-(2,2-Dichlorovinyl)-2,2-dimethylcyclopropane carboxylic acid

In GLM analyses, a 50% increase of home distance to floriculture greenhouses was associated with a 2.06% (95% confidence interval [CI]: 0.29%, 3.87%) increase in TCPy concentration (Table 3; Fig. 2a); no statistically significant associations were observed with any other urinary pesticide metabolite. There was evidence of a curvilinear association (p<0.05) between home distance and PNP concentration (Table 3).

**Figure 2.**
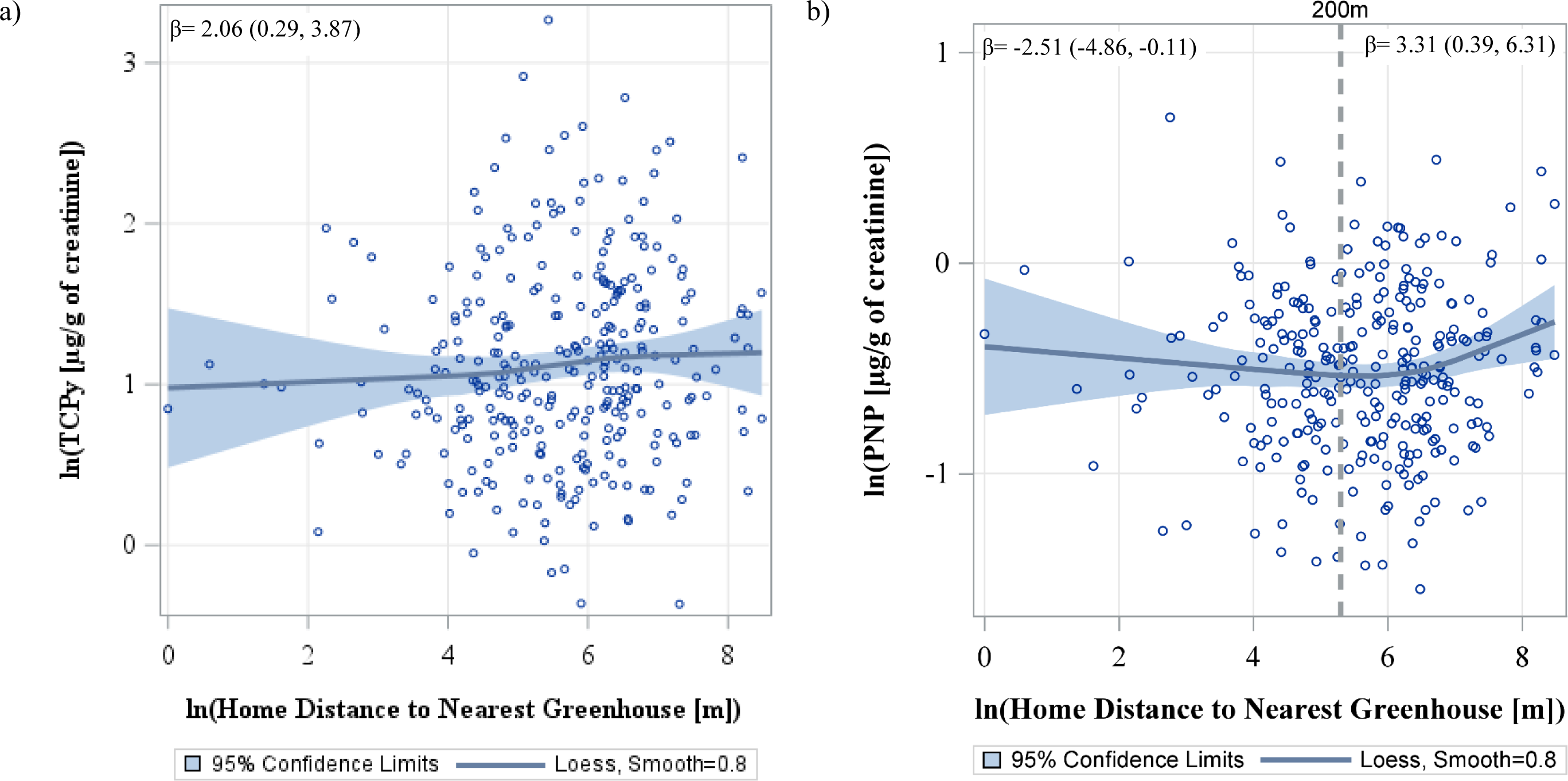
LOESS graph of the adjusted associations between home proximity to the nearest greenhouse, with PNP and TCPy concentration. Each data point represents the adjusted least squares of ln(PNP) or ln(TCPy) concentration for 350 ranks of ln(home distance to the nearest greenhouse [m]). The models adjusted for age, height-for-age z-score, BMI-for-age z-score, race, gender, monthly income, parental education, living with an agricultural or flower worker. The blue line represents the LOESS line, while the shaded blue regions are the 95% confidence intervals for the LOESS line. For Figure 1a, the β estimate is the linear association between home distance to the nearest greenhouses and TCPy (see Table 2) For Figure 1b, a 200m reference line (equal to a y value of 5.3) is shown to to represent the cut-off point used in our stratified analysis (see Table 3), and the respective β estimate for each strata is shown. The models were adjusted for age, height-for-age z-score, BMI-for-age z-score, race, gender, monthly income, parental education, living with an agricultural or flower worker PNP= para-Nitrophenol, TCPy= 3,5,6-Trichloro-2-pyridinol, m= meters, µg=microgram, g=gram

Compared to the linear regression estimates (Table 3), the associations strengthened for all metabolites when Tobit regression was conducted (Table S3), however, the associations were not statistically significant. This is with the exception of *trans-*DCCA whose association remained the same.

**Table 3:**
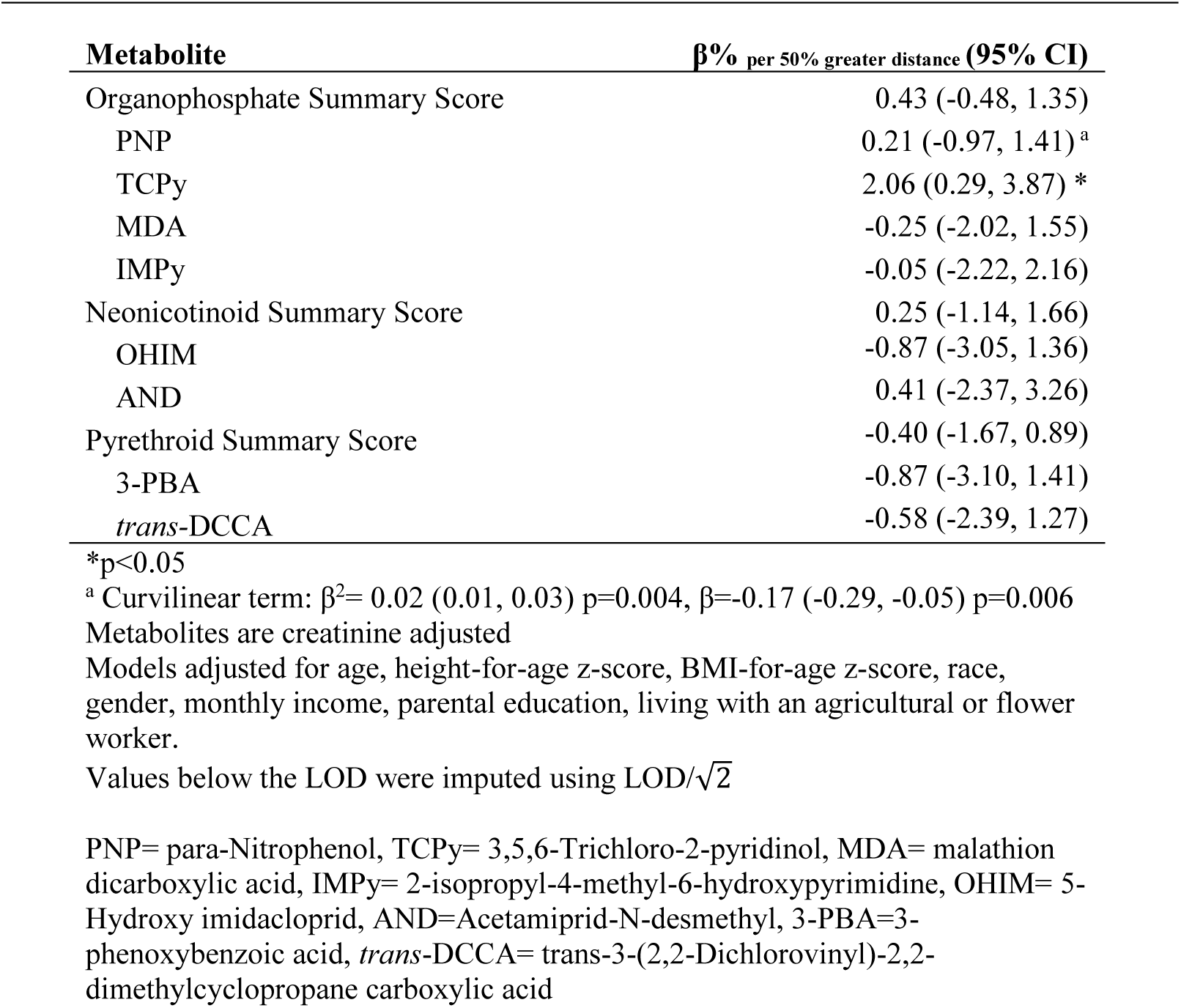
Percent difference of metabolite concentration per 50% increase in residential distance to the nearest floricultural greenhouse (β%_per 50% greater distance_ (95% CI)).

### Stratification by proximity

We stratified associations between home distance and metabolite concentrations using a 200m cut point (Table 4); this cut-off was chosen based on sensitivity analysis (Tables S1). A potential threshold effect for PNP was identified (Table 3), as there was a negative association amongst those who lived within 200m of a greenhouse (βper 50% increase=-2.51% [-4.86%, -0.11%]), which was positive amongst those who lived outside of 200m of a greenhouse (βper 50% increase=3.31% [0.39%, 6.31%]). These slope differences could be visualized in the LOESS graph (Fig. 2b). Similar threshold effects were observed for the organophosphate summary score, IMPy, the neonicotinoid summary score, and AND; however, none of the associations reached statistical significance.

**Table 4.**
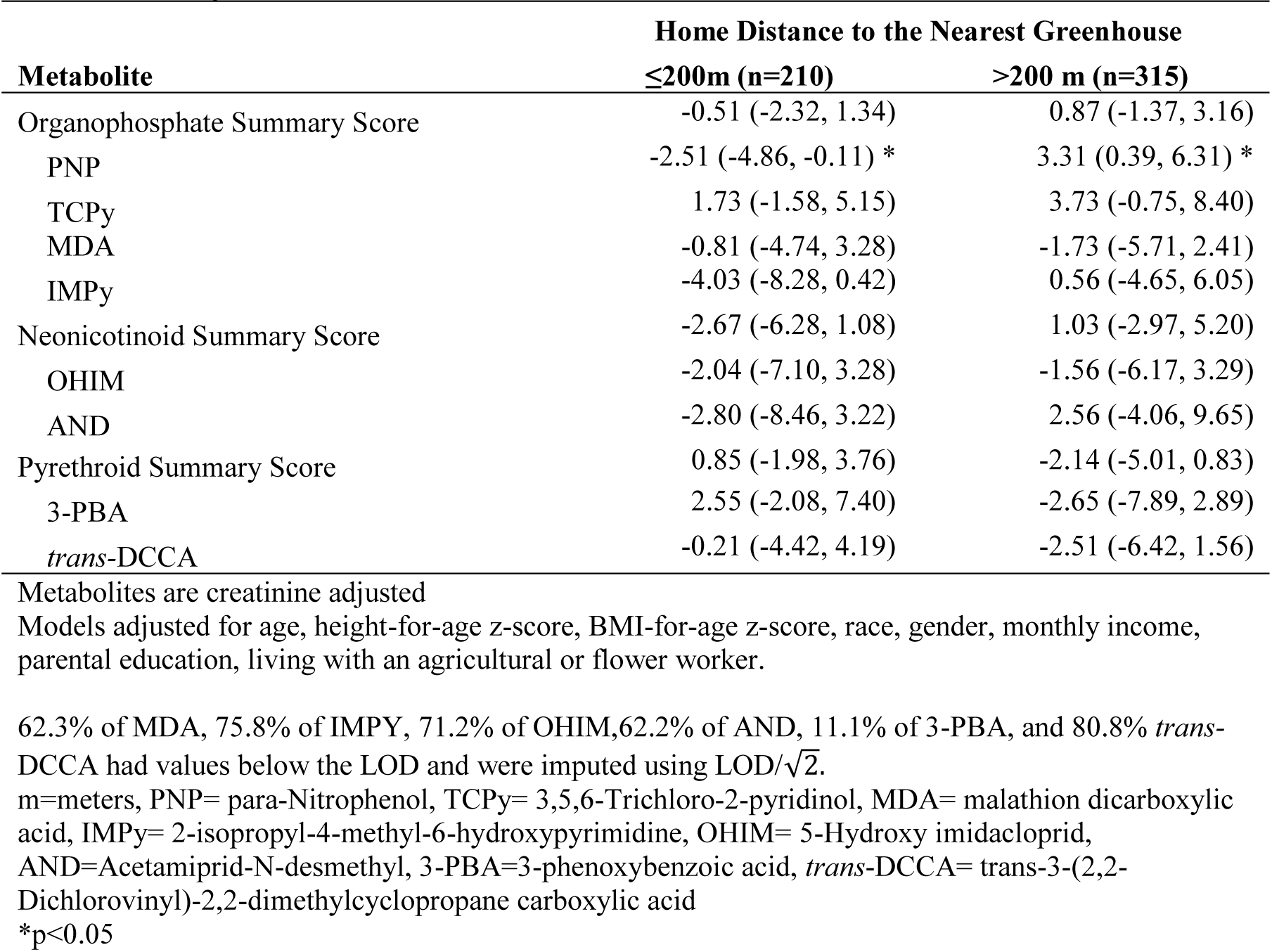
Percent difference of metabolite concentration per 50% increase of residential distance to the nearest floricultural greenhouse (β%_per 50% greater distance_ (95% CI)), stratified by residential distance to greenhouse of 200m.

| Metabolite | Home Distance to the Nearest Greenhouse |  |
| --- | --- | --- |
| | $\leq 200\text{m}$ (n=210) | $> 200\text{ m}$ (n=315) |
| Organophosphate Summary Score | -0.51 (-2.32, 1.34) | 0.87 (-1.37, 3.16) |
| PNP | -2.51 (-4.86, -0.11) * | 3.31 (0.39, 6.31) * |
| TCPy | 1.73 (-1.58, 5.15) | 3.73 (-0.75, 8.40) |
| MDA | -0.81 (-4.74, 3.28) | -1.73 (-5.71, 2.41) |
| IMPy | -4.03 (-8.28, 0.42) | 0.56 (-4.65, 6.05) |
| Neonicotinoid Summary Score | -2.67 (-6.28, 1.08) | 1.03 (-2.97, 5.20) |
| OHIM | -2.04 (-7.10, 3.28) | -1.56 (-6.17, 3.29) |
| AND | -2.80 (-8.46, 3.22) | 2.56 (-4.06, 9.65) |
| Pyrethroid Summary Score | 0.85 (-1.98, 3.76) | -2.14 (-5.01, 0.83) |
| 3-PBA | 2.55 (-2.08, 7.40) | -2.65 (-7.89, 2.89) |
| *trans*-DCCA | -0.21 (-4.42, 4.19) | -2.51 (-6.42, 1.56) |

Metabolites are creatinine adjusted
Models adjusted for age, height-for-age z-score, BMI-for-age z-score, race, gender, monthly income, parental education, living with an agricultural or flower worker.
62.3% of MDA, 75.8% of IMPY, 71.2% of OHIM, 62.2% of AND, 11.1% of 3-PBA, and 80.8% *trans*-DCCA had values below the LOD and were imputed using $\text{LOD}/\sqrt{2}$ .
\* $p < 0.05$

**Table 5.** Percent difference of metabolite concentration for every 50% increase in surface area within 150m from homes (β% _per 50% greater area_ [95%CI]).

| Metabolite | $\beta\%$ per 50% greater area (95% CI) |
| --- | --- |
| Organophosphate Summary Score | -0.25 (-0.58, 0.08) |
| PNP | 0.06 (-0.36, 0.50) |
| TCPy | -0.66 (-1.28, -0.02) * |
| MDA | -0.30 (-0.94, 0.35) |
| IMPy | 0.01 (-0.78, 0.81) |
| Neonicotinoid Summary Score | 0.19 (-0.44, 0.82) |
| OHIM | 0.13 (-0.68, 0.95) |
| AND | 0.47 (-0.56, 1.50) |
| Pyrethroid Summary Score | 0.11 (-0.35, 0.58) |
| 3-PBA | 0.39 (-0.44, 1.22) |
| *trans*-DCCA | 0.08 (-0.58, 0.75) |

Metabolites outcomes are creatinine adjusted
Models adjusted for age, height-for-age z-score, BMI-for-age z-score, race, gender, monthly income, parental education, living with an agricultural or flower worker.
62.3% of MDA, 75.8% of IMPY, 71.2% of OHIM, 62.2% of AND, 11.1% of 3-PBA, and 80.8% *trans*-DCCA had values below the LOD and were imputed using LOD/ $\sqrt{2}$ .
\*p<0.05

### Greenhouse surface area and urinary metabolite concentrations

Floricultural surface area near homes was negatively associated with TCPy concentrations at the 150m buffer size (β=-0.66% [-1.28%, -0.02%], Table 4). As expected, smaller buffers were associated with stronger associations with TCPy compared to larger buffers, except for the 750m buffer (Table S4). No other statistically significant associations were observed with other metabolites. Using Tobit regression, the associations between floricultural surface area with IMPY, MDA, OHIM, AND, and 3-PBA (Table S3) were stronger compared to linear regression estimates but were not statistically significant for either.

Using Moran’s I, we identified that the linear regression residuals were clustered for all metabolite models (Table S3); the hotspot (Fig 3 and 4) analyses indicated that the location of the hot and cold spots varied by metabolite. For organophosphates, we observed hotspots for TCPy (Fig.3a), MDA (Fig. 3b), PNP (Fig. 3c), and IMPy (Fig. 3d) parish of Tupigachi, along the eastern border of the county. MDA and PNP had a cold spot in central Malchingui, while TCPy and PNP had a cold spot in Tabacundo. The location of hot and cold spots varied more for the pyrethroids and neonicotinoids, compared to the organophosphate metabolites. Beginning with the neonicotinoids, OHIM (Fig. 3a) had a hotspot in central Tocachi, southern La Esperanza, and near the greenhouses in the southern region of Tupigachi. While AND (Fig. 4b) had a hotspot in Malchingui and La Esperanza, but a cold spot in Tupigachi. For the pyrethroids, 3-PBA (Fig. 4c) and *trans*-DCCA (Fig. 4d) both had hotspots located in central to southern La Esperanza, and southern Tupigachi. The cold spot locations varied, as 3-PBA had a small cold spot in central eastern Tabacundo, while *trans-*DCCA had one in Malchingui and in the border of Tabacundo and Tupigachi. GWR was conducted for TCPy and PNP, as these were the only two significant associations we identified in the overall and stratified linear regression results (Figure 5). For TCPy, we found negative associations with distance in the Malchingui region (βrange=-0.22-0). There were positive associations between a 500m increase in distance and TCPy (βrange=0.01-0.54) in all parishes except Malchingui. The smallest positive β values were present in Tupigachi (βrange=0.01-0.34) and Malchingui (βrange=0.01-0.16), while the largest positive estimates were in Tocachi (βrange=0.17-0.54) and La Esperanza (βrange=0.17-0.54). The largest standard errors were in Tocachi and La Esperanza. For PNP, there were negative associations with a 500 m increase of distance in Tocachi, La Esperanza, and the southern region of Tabacundo. Positive associations between distance (per 500m) and PNP concentration were present in Malchingui and Tupigachi.

**Figure 3.**
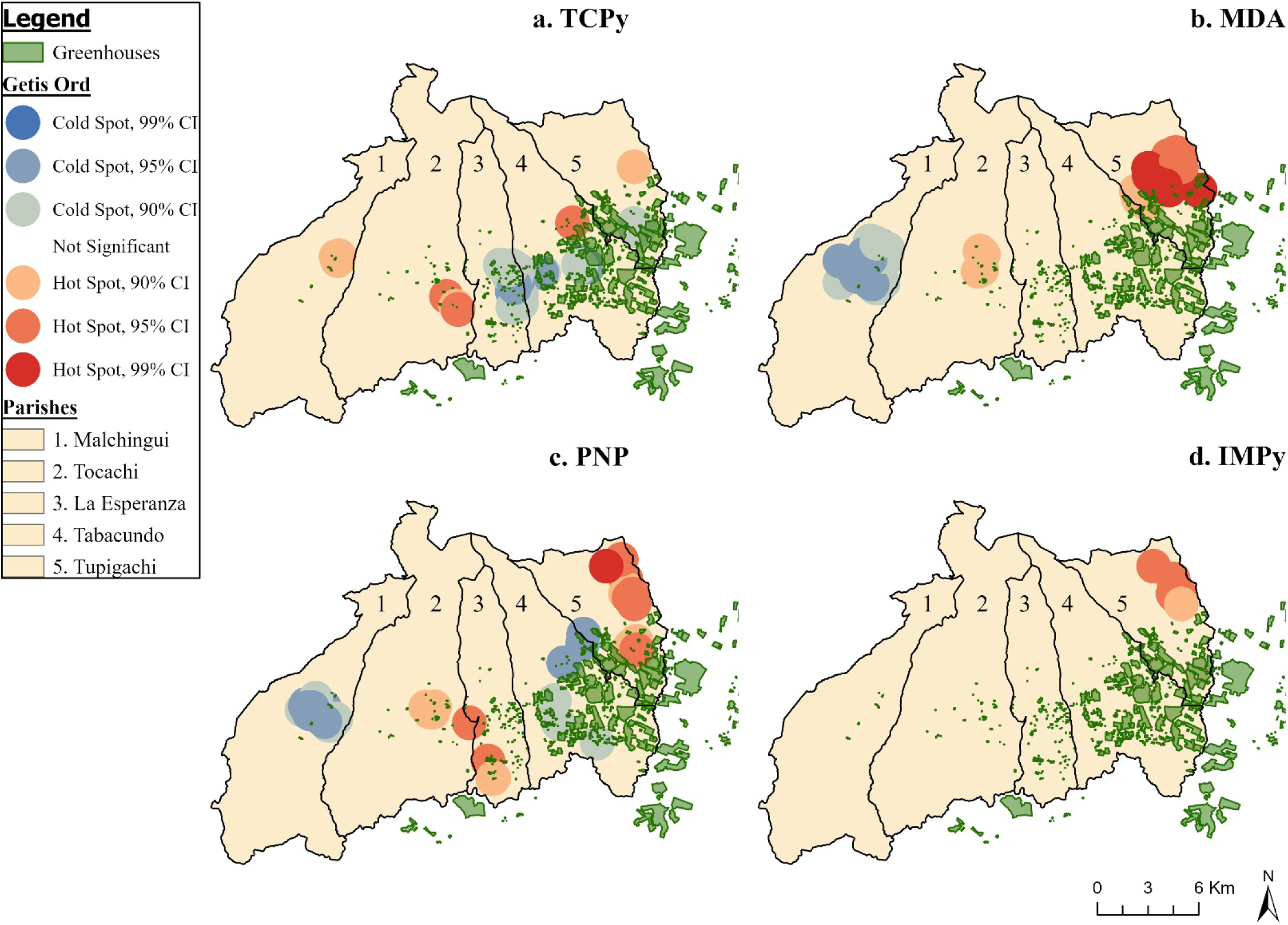
Hot Spot Analysis (Getis-Ord Gi*) of organophosphate metabolite concentrations in Pedro Moncayo, Ecuador. PNP= para-Nitrophenol, TCPy= 3,5,6-Trichloro-2-pyridinol, MDA= malathion dicarboxylic acid, IMPy= 2-isopropyl-4-methyl-6-hydroxypyrimidine, CI=Confidence Interval

**Figure 4.**
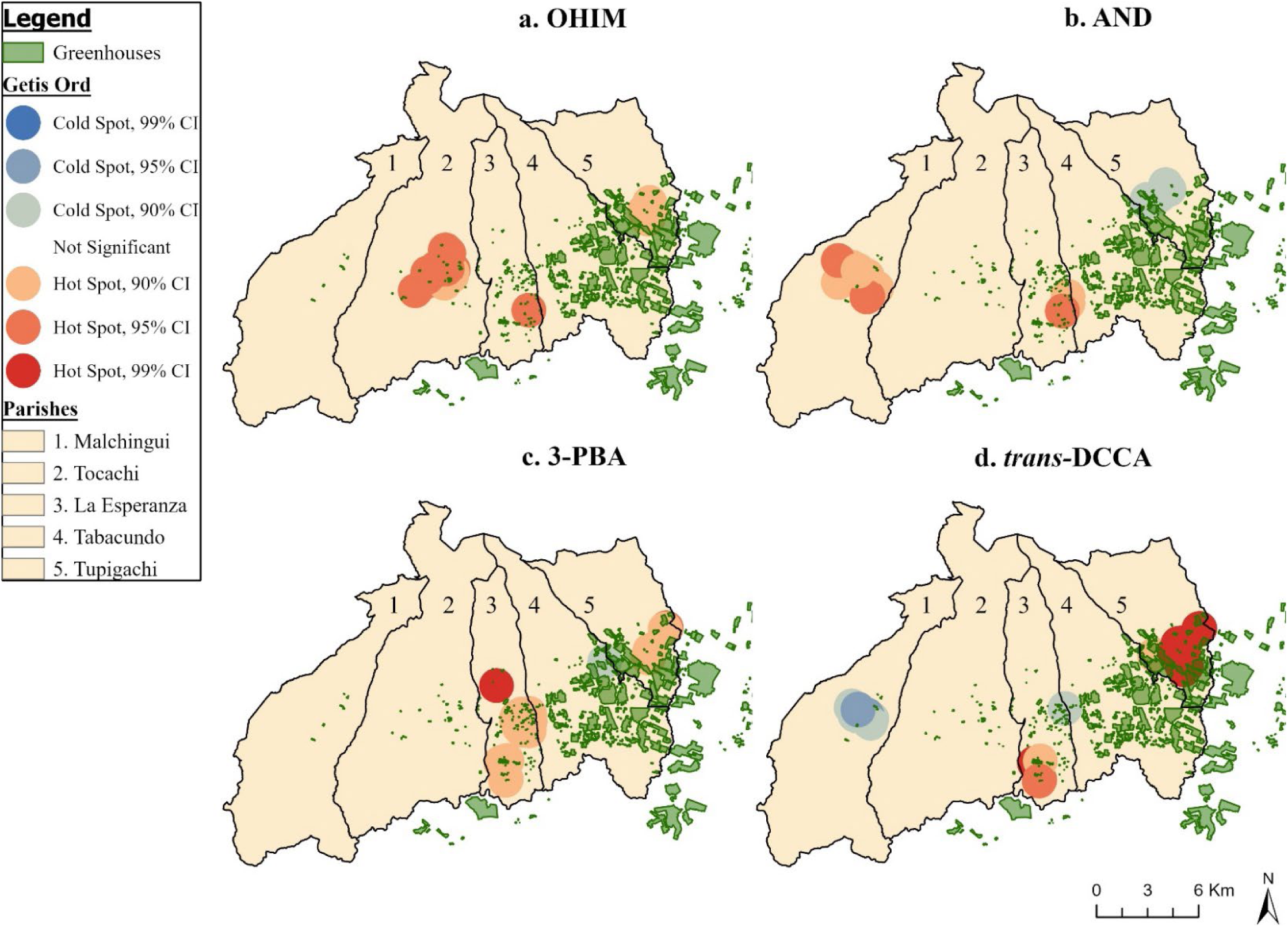
Hot Spot Analysis (Getis-Ord Gi*) of pyrethroid (3-PBA & trans-DCCA) and neonicotinoid (OHIM & AND) metabolite concentrations in Pedro Moncayo, Ecuador. All non-statistically significant hot and cold spots have been masked to preserve de-identification of participant homes. m=meters, OHIM= 5-Hydroxy imidacloprid, AND=Acetamiprid-N-desmethyl, 3-PBA=3-phenoxybenzoic acid, trans-DCCA= trans-3-(2,2-Dichlorovinyl)-2,2-dimethylcyclopropane carboxylic acid, CI=Confidence Interval.

**Figure 5.**
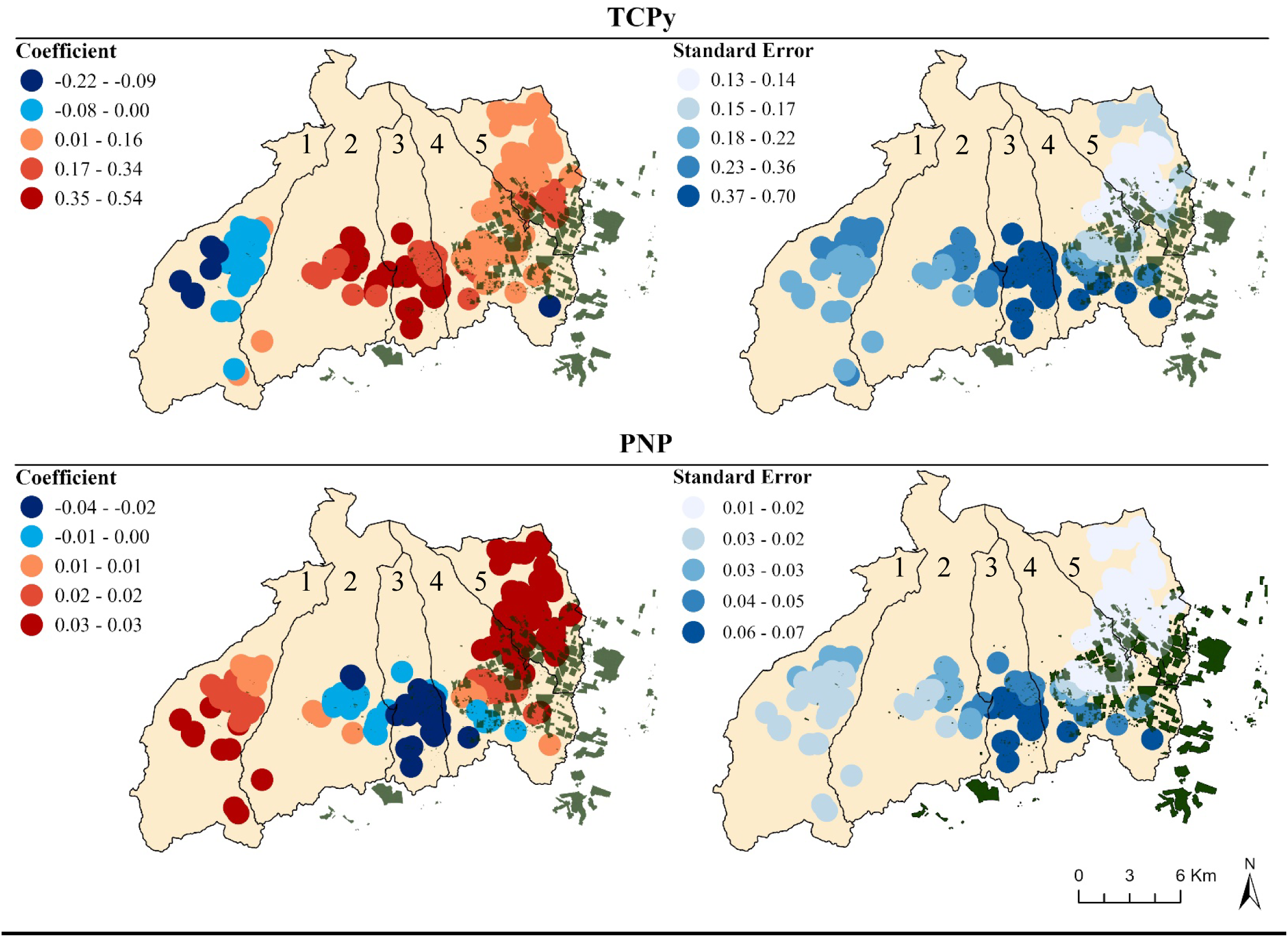
Distribution of geospatially weighted regression coefficients and standard error, for the associations between a 500m increase in distance to floriculture with TCPy and PNP concentration. 1=Malchingui, 2=Tocachi, 3=La Esperanza, 4=Tabacundo, 5=Tupigachi GWR points have radii of 1km to preserve de-identification of participant homes. Models were adjusted for age, BMI-for-age z-score, average monthly salary (USD), average parental education, and cohabitation with a floricultural or agricultural worker. m=meters, PNP= para-Nitrophenol, TCPy= 3,5,6-Trichloro-2-pyridinol

## 4. Discussion

Our findings suggest that greenhouse floriculture is a source of off-target drift for the organophosphates parathion or methyl parathion (parent chemicals of PNP) and diazinon (parent chemical of IMPy), and for the neonicotinoid imidacloprid (parent chemical of OHIM). During this same study period, however, floriculture did not appear to be a source of exposure of the organophosphate chlorpyrifos (parent chemical of TCPy). To our knowledge, this study is the first to classify whether proximity to greenhouse agriculture influences the presence of urinary pesticide metabolites in adolescents. For PNP, we observed a significant curvilinear association with residential distance to floriculture greenhouses, in which home distance was negatively associated with PNP concentration among participants living within 200m of a greenhouse, while the association was positive for those living at greater distances. The same pattern was observed for IMPy. OHIM concentrations were found to be higher for participants living within 200m of a greenhouse, compared to those who lived outside of 200m. TCPy concentration was positively associated with home distance to the nearest floricultural greenhouse, and increasing surface area of greenhouses were associated with lower TCPy concentration. Thus, we found that associations between residential distance to greenhouses and urinary pesticide metabolites differed by metabolite type.

The negative associations we observed of residential proximity to floricultural greenhouses with PNP (and borderline non-significant with IMPy) for individuals living within 200m of such greenhouses is concordant with previous findings within the ESPINA study, in which a positive association between residential proximity to floricultural greenhouses and AChE activity was observed among participants who lived within 275m from crops^67^ (lower AChE activity reflects greater exposure to organophosphates and carbamates). The potential threshold effect observed may be due to reduced pesticide exposure following certain distances from crops. For example, air concentrations of pesticides were found to decline steeply after 250m of distance from tulip fields following pesticide application in the Netherlands.^59^ Contrary to the PNP results, farther home distance to floricultural greenhouses and less floricultural surface areas near homes had higher TCPy concentrations, overall and across the 200m strata. These findings suggest that chlorpyrifos, the parent chemical of TCPy, may not have been used in floriculture in the weeks prior to the examination of ESPINA participants or that there may be non-floricultural sources of pesticide exposure in this population. Pedro Moncayo County has a considerable year-round production of non-floricultural agricultural products such as wheat, barley, corn, legumes, and berries, on which pesticides are often sprayed. Further research to identify the sources of chlorpyrifos exposures in this population is warranted.

Although we identified few linear relationships between home distance to floriculture and home surface area of floriculture with metabolite concentrations, this may have been due to spatial clustering and regional variation in the associations. The proportion of the total area areas devoted to floricultural greenhouses varied substantially by parish: Malchingui (0.05%), Tupigachi (0.12%), La Esperanza (1.12%), Tabacundo (13.1%) and Tupigachi (12.4%). A common hotspot for organophosphate exposure in our analyses occurred in Tupigachi, for pyrethroids it occurred in La Esperanza and Tupigachi, and for neonicotinoids in Malchingui and Tocachi. Using GWR we identified that there were negative associations between home distance to floriculture and TCPy concentrations in Malchingui, while we identified positive associations in the four remaining parishes. For PNP, home distance to floriculture was negatively associated with PNP concentration in Tocachi, La Esperanza, and parts of Tabacundo, but was positively associated everywhere else. The diversity of GWR estimates demonstrates that there may be different patterns of pesticide use exposure across the five parishes of Pedro Moncayo, explaining limited significance in the global regression results.

In the hotspot analysis for neonicotinoids OHIM and AND, hot spots were located in Malchingui and Tocachi, two parishes in western Pedro Moncayo that are less densely occupied by floriculture and where considerable non-floricultural agricultural production exists. In 2008, it was estimated that 58.1% of the total area of Pedro Moncayo County was occupied by agriculture^68^ and floriculture occupied 2% of the geographic area (656 ha).^43^ By 2016, floricultural crops more than doubled, covering 4.47% (1495 ha) of land.^45^ Thus, it is plausible that pesticide use in non-floricultural agriculture production is contributing to the neonicotinoid exposure in our cohort. OHIM, however, also had a hotspot in Tupigachi, and had higher ls-mean concentrations for participants living within 200m of floricultural greenhouses, indicating that pesticide drift of imidacloprid, OHIM’s parent chemical, may be coming from select greenhouses in this region.

The hot spot analysis identified that there are localized differences in pesticide exposure, with differing patterns observed between pesticide types (organophosphates, neonicotinoids, or pyrethroids), and across the metabolites themselves. This may be due to differences in pesticide use across the floricultural and agricultural industry. The intensity of pesticide use, a metric consisting of the amount and number of active ingredients applied to crops,^69^ generally varies by the target crops (i.e., rose, corn, wheat, etc.), but may also vary across the companies and individual farm holders that own the cropland. Importation regulations on flower-based food products and biosafety limit the amount of pesticide residues that can be present, but there is a lack of maximum residue limits for import of ornamental flowers which leads to limited restriction of pesticide use across the industry.^39,70^ In Belgium where 36% of flower imports come from Ecuador, 60 active pesticide residues (primarily fungicides and insecticides) were detected on roses imported from Ecuador (n=9 rose samples) obtained from 25 different florists.^71^ The roses from Ecuador had the greatest number of active pesticide residues (n=60 active pesticide residues), compared to roses sampled from 9 other countries (nrange=24-54 active pesticide residues).^71^ In a review of 81 studies about pesticides use in global flower production, organophosphates were the most frequently detected (n=34), pyrethroids were fourth (n=13), while neonicotinoids were sixth out of 10 measured insecticide classifications reported.^39^ This pattern was seen for the organophosphate and pyrethroids metabolites, as hotspots occurred in parishes with higher density of floricultural presence. Differences in hotspots between the organophosphate metabolites may be due to different pesticide application practices by different companies, as there are 148 unique rose companies present in Pedro Moncayo.^72^, As urinary metabolites measure generally measure recent exposure to pesticides, variations in pesticide use across the crop types and companies could have resulted in the varying distribution of hot and cold spots.

The distribution of hotspots, and differences in GWR coefficients might also be due to pesticide exposure pathways other than pesticide drift, such as other para-occupational sources like take-home pesticide exposures from agricultural workers. Take-home pesticide exposure pathways are a well-documented risk for increased pesticide exposure in family members of agricultural workers,^60,73^ and we previously identified that children’s cohabitation with an agricultural worker was associated with lower AChE activity in this study population, marking greater pesticide exposure, compared to children who did not live with agricultural workers.^1^ While our analyses controlled for participant cohabitation with floricultural or other agricultural workers, it is possible that a marginal amount of residual confounding may be present considering that floricultural workers may tend to live closer to floricultural crops.

The strengths of this study include a generally large sample size with a considerable distribution of participants living near and far from floricultural greenhouses. Additionally, our analyses include specific urinary metabolites for the major classes of insecticides used worldwide, which reduces misclassification of our main outcomes. Limitations of our study include not accounting for wind patterns or information regarding the distribution of non-floricultural agriculture in Pedro Moncayo, which would allow us to better characterize the sources of pesticide drift. There are no maps of non-floricultural agriculture, like corn, in Pedro

Moncayo for 2016. This region has a high presence of smallholder farming (10 hectares or less)^74^, thus drawing agricultural fields through visual inspection of satellite imagery would have introduced misclassification bias. Incorporating remote sensing methods to map agriculture would allow characterization of pesticide drift from other agriculture. Furthermore, urinary pesticide metabolite concentrations represent recent exposure to pesticides, as they are metabolized quickly (∼72 hours); therefore, including multiple metabolite measurements would reduce exposure misclassification and improve the precision of our results. Our study was unable to control for potential pesticide exposure by other exposure pathways, like diet, but we were able to control for para-occupational exposures. Another weakness is that our model assumes participants are static. Geospatial analyses, such as those presented in this study, could also be strengthened by individual based models of exposure that track participants using smartphones or global positioning system instruments to better assess movement and duration of proximity to known exposure sites.^75–77^

## 5. Conclusion

We observed negative associations between residential distance to greenhouses (within 200m) with urinary concentrations of PNP (metabolite of parathion), MDA (metabolite of malathion) and OHIM (metabolite of imidacloprid) in adolescents in this population and study period. This suggests that these pesticides can drift from floricultural greenhouses and reach children living nearby. We also observed positive associations of between residential distance to greenhouses and TCPy (metabolite of chlorpyrifos) concentrations, which suggest that exposure to this insecticide during the present study period may stem from non-floricultural open-air crops, such as corn, wheat, strawberries, etc. Our analyses of this study period suggest that parathion (PNP parent chemical), malathion (MDA parent chemical) and imidacloprid (OHIM parent chemical) exposure of adolescents in this population may stem from pesticide drift from floricultural greenhouses, while chlorpyrifos (TCPy parent chemical) exposure may stem from open-air agriculture, such as corn, wheat, or strawberries. This study also found localized differences across the five parishes of Pedro Moncayo County. Organophosphates and pyrethroids were found to have hotspots in regions with greater floricultural surface area, while neonicotinoids had hotspots in regions where there might be a higher presence of non-floricultural agriculture. Our findings demonstrates that there might be differential patterns of pesticide use between floriculture and agriculture in this region, as intensity of pesticide use varies by crop type. Seasonality for these crops could affect pesticide use as well, as application practices might vary between rose and non-rose crops. Given the negative health consequences that arise from pesticide exposure, this study highlights the need to assess spatial variations in pesticide exposure across seasons, understand pesticide application practices across crops, and to mitigate pesticide exposures of populations living near pesticide spray sites.

## Data Availability

All data produced in the present study are available upon reasonable request to the authors

https://knit.ucsd.edu/espina/

## Acknowledgments

The ESPINA study received funding from the National Institute of Occupational Safety and Health (1R36OH009402) and the National Institute of Environmental Health Sciences (R01ES025792, R01ES030378, R21ES026084, U2CES026560, P30ES019776). We thank Fundación Cimas del Ecuador, the Parish Governments of Pedro Moncayo County, community members of Pedro Moncayo and the Education District of Pichincha-Cayambe-Pedro Moncayo counties for their support on this project. B. Chronister was supported by the National Institute of Mental Health (5T32MH122376). Funding was provided by NIEHS K01ES031697 to Dr. Kayser that helped bring this research to publication.

---

**Table S1.**
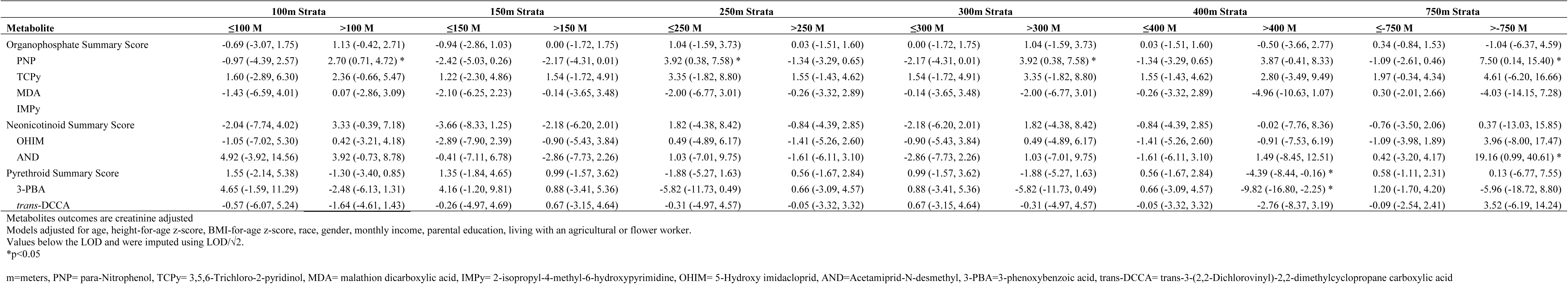
Percent difference (β% [95%CI]) of metabolite concentration for every 50% increase in distance from the home to the nearest greenhouse using a 100m, 150m, 250m, 300m, 400m, and 750m strata.

**Table S2.** Percent difference of metabolite concentration per 50% increase in residential distance to the nearest floricultural greenhouse (β%_per 50% greater distance_ (95% CI)) using Tobit regression.

| Metabolite | $\beta\%$ per 50% greater distance (95% CI) | Standard Error | P-value |
| --- | --- | --- | --- |
| IMPy | -1.83 | 3.18 | 0.56 |
| MDA | -1.12 | 1.65 | 0.49 |
| OHIM | -1.91 | 2.67 | 0.46 |
| AND | 0.49 | 2.98 | 0.87 |
| 3-PBA | -0.96 | 1.21 | 0.43 |
| *trans*-DCCA | -0.58 | 0.93 | 0.53 |

\* $p < 0.05$
Models adjusted for age, height-for-age z-score, BMI-for-age z-score, race, gender, monthly income, parental education, living with an agricultural or flower worker.
MDA= malathion dicarboxylic acid, IMPy= 2-isopropyl-4-methyl-6-hydroxypyrimidine, OHIM= 5-Hydroxy imidacloprid, AND=Acetamiprid-N-desmethyl, 3-PBA=3-phenoxybenzoic acid, *trans*-DCCA= *trans*-3-(2,2-Dichlorovinyl)-2,2-dimethylcyclopropane carboxylic acid
The left bound censoring value was determined by taking the LOD, dividing by creatinine, and ln-transforming the value.

**Table S3.** Percent difference of metabolite concentration for every 50% increase in surface area within 150m from homes (β% _per 50% greater area_ [95%CI]) using Tobit regression.

| Metabolite | $\beta\%$ per 50% greater area<br>(95% CI) | Standard Error | P-value |
| --- | --- | --- | --- |
| IMPy | 1.20 | 1.11 | 0.28 |
| MDA | -0.44 | 0.58 | 0.45 |
| OHIM | 0.25 | 0.95 | 0.79 |
| AND | 1.04 | 1.06 | 0.33 |
| 3-PBA | 0.41 | 0.44 | 0.35 |
| *trans-DCCA* | 0.08 | 0.34 | 0.81 |

\*p<0.05
Models adjusted for age, height-for-age z-score, BMI-for-age z-score, race, gender, monthly income, parental education, living with an agricultural or flower worker.
PNP= para-Nitrophenol, TCPy= 3,5,6-Trichloro-2-pyridinol, MDA= malathion dicarboxylic acid, IMPy= 2-isopropyl-4-methyl-6-hydroxypyrimidine, OHIM= 5-Hydroxy imidacloprid, AND=Acetamiprid-N-desmethyl, 3-PBA=3-phenoxybenzoic acid, trans-DCCA= trans-3-(2,2-Dichlorovinyl)-2,2-dimethylcyclopropane carboxylic acid
The left bound censoring value was determined by taking the LOD, dividing by creatinine, and ln-transforming the value.

**Table S4.** Percent change (β% [95%CI]) of metabolite concentration for every 50% increase in surface area concentration within a 100m, 200m, 300m, 500m, and 750m buffer size.

| Metabolite | 100m | 200m | 300m | 500m | 750m |
| --- | --- | --- | --- | --- | --- |
| Organophosphate Summary Score | -0.10 (-0.50, 0.31) | -0.18 (-0.47, 0.12) | -0.13 (-0.39, 0.14) | -0.15 (-0.39, 0.10) | -0.15 (-0.42, 0.13) |
| PNP | 0.32 (-0.20, 0.86) | -0.05 (-0.43, 0.34) | -0.12 (-0.46, 0.22) | -0.17 (-0.49, 0.15) | -0.31 (-0.66, 0.04) |
| TCPy | -0.71 (-1.48, 0.07) | -0.49 (-1.06, 0.08) | -0.45 (-0.95, 0.06) | -0.43 (-0.90, 0.05) | -0.53 (-1.05, -0.002) * |
| MDA | 0.01 (-0.79, 0.81) | -0.16 (-0.74, 0.42) | -0.05 (-0.57, 0.47) | -0.05 (-0.53, 0.44) | 0.08 (-0.45, 0.62) |
| IMPy | 0.65 (-0.32, 1.64) | -0.20 (-0.91, 0.51) | -0.09 (-0.72, 0.54) | -0.03 (-0.62, 0.57) | -0.01 (-0.66, 0.65) |
| Neonicotinoid Summary Score | 0.57 (-0.20, 1.34) | -0.14 (-0.59, 0.31) | -0.03 (-0.53, 0.47) | -0.01 (-0.48, 0.45) | 0.17 (-0.35, 0.68) |
| OHIM | 0.62 (-0.39, 1.63) | -0.04 (-0.95, 0.88) | 0.35 (-0.30, 1.00) | 0.23 (-0.38, 0.84) | 0.33 (-0.33, 1.01) |
| AND | 0.89 (-0.37, 2.16) | -0.24 (-0.72, 0.23) | -0.25 (-1.06, 0.56) | -0.24 (-1.00, 0.53) | 0.05 (-0.79, 0.90) |
| Pyrethroid Summary Score | 0.03 (-0.54, 0.61) | 0.04 (-0.38, 0.45) | 0.14 (-0.23, 0.51) | 0.14 (-0.21, 0.49) | 0.32 (-0.07, 0.70) |
| 3-PBA | 0.20 (-0.81, 1.23) | 0.29 (-0.45, 1.03) | 0.08 (-0.58, 0.73) | 0.12 (-0.49, 0.74) | 0.53 (-0.15, 1.21) |
| *trans*-DCCA | -0.04 (-0.85, 0.79) | 0.02 (-0.58, 0.62) | 0.30 (-0.23, 0.83) | 0.25 (-0.25, 0.75) | 0.30 (-0.25, 0.86) |

Metabolites outcomes are creatinine adjusted
Models adjusted for age, height-for-age z-score, BMI-for-age z-score, race, gender, monthly income, parental education, living with an agricultural or flower worker.
62.3% of MDA, 75.8% of IMPY, 71.2% of OHIM, 62.2% of AND, 11.1% of 3-PBA, and 80.8% *trans*-DCCA had values below the LOD and were imputed using LOD/ $\sqrt{2}$ .
\*p<0.05

**Table S5.** Moran’s I for linear regression residuals which assessed the association between home distance to the nearest greenhouse and urinary pesticide metabolite concentration.

|  | OLS Residuals |  |  |
| --- | --- | --- | --- |
|  | Moran I | z-score | p-value |
| Organophosphate Summary Score | 0.14 | 3.63 | <0.001 |
| PNP | 0.12 | 3.13 | <0.001 |
| TCPY | 0.19 | 5.20 | <0.001 |
| MDA | 0.13 | 3.57 | <0.001 |
| IMPY | 0.18 | 4.79 | <0.001 |
| Neonicotinoid Summary Score | 0.09 | 2.32 | 0.02 |
| OHIM | 0.11 | 3.10 | <0.001 |
| AND | 0.11 | 2.80 | 0.01 |
| Pyrethroid Summary Score | 0.16 | 4.21 | <0.001 |
| 3-PBA | 0.18 | 4.71 | <0.001 |
| *trans*-DCCA | 0.12 | 3.15 | <0.001 |

PNP= para-Nitrophenol, TCPy= 3,5,6-Trichloro-2-pyridinol, MDA= malathion dicarboxylic acid, IMPy= 2-isopropyl-4-methyl-6-hydroxypyrimidine, OHIM= 5-Hydroxy imidacloprid, AND=Acetamiprid-N-desmethyl, 3-PBA=3-phenoxybenzoic acid, *trans*-DCCA= *trans*-3-(2,2-Dichlorovinyl)-2,2-dimethylcyclopropane carboxylic acid

